# Adherence to the EAT-Lancet diet and risk of incident type 2 diabetes: the Danish Diet, Cancer and Health cohort

**DOI:** 10.1101/2021.12.16.21267887

**Authors:** Fie Langmann, Daniel B. Ibsen, Anne Tjønneland, Anja Olsen, Kim Overvad, Christina C. Dahm

## Abstract

**Objective:** In 2019 the EAT-Lancet Commission proposed a dietary pattern, defined to be globally environmentally sustainable, but untested directly in population studies with regards to health. We investigated adherence to the EAT-Lancet diet and risk of incident type 2 diabetes in a Danish setting.

**Research design and methods:** In total, 54,232 participants aged 50-64 years at inclusion (1993-1997) with no previous cancer or diabetes diagnoses were included. Dietary data were collected using a validated 192-item food frequency questionnaire, and scored 0 (non-adherence) or 1 (adherence) point for each of the 14 dietary components of the EAT-Lancet diet (range 0-14 points). Incident type 2 diabetes cases were identified using the Danish National Diabetes Register. Hazard ratios and 95% confidence intervals (CI) were estimated using multivariable-adjusted Cox proportional hazards models.

**Results:** During follow-up (median 15 years), 7130 participants developed type 2 diabetes. There was a 22% (95% CI: 14%; 29%) lower risk of type 2 diabetes among those with the greatest adherence to the EAT-Lancet diet (11-14 points) compared to those with the lowest adherence (0-7 points). After further adjusting for potential mediators, the corresponding risk was 17% (95% CI: 8%; 24%) lower.

**Conclusion:** Adherence to the EAT-Lancet diet was associated with a lower risk of developing type 2 diabetes in a middle-aged Danish population.

## Introduction

In 2019, 9.3% of the world’s population were estimated to be affected by type 2 diabetes (1). Type 2 diabetes entails elevated risk of debilitating complications such as cardiovascular disease, and causes more than one million deaths yearly (2, 3). Of modifiable risk factors, dietary patterns have shown to play a key role in prevention of type 2 diabetes (3).

Dietary patterns with lower consumption or exclusion of meat, such as plant-based, flexitarian, and pescatarian diets, have been associated with lower risk of type 2 diabetes (4). Higher intakes of plant-based foods rich in vitamins, minerals, and antioxidants, and lower intake of red and processed meats might underlie the protective association (4-7). However, complete exclusion of animal products can also exclude beneficial food groups such as dairy products and fish that potentially prevent type 2 diabetes, and not all plant-based foods are equally healthy (8-10). High consumption of refined grains, starches, and sugars are associated with a higher risk of type 2 diabetes (9, 10). Thus, there are opposing effects of different components of plant-based dietary patterns on the risk of type 2 diabetes (11).

Our diets not only play a critical role in human health. Growing attention is currently being devoted to integrate human health and planetary sustainability in a common global agenda for food system transformation (12). In 2019, the EAT-Lancet Commission defined a universal reference diet to promote population and planetary health. The diet bears some resemblance to meat-restrictive diets such as vegetarian or vegan diets, albeit with low meat consumption. Since dairy and fish are included in the EAT-Lancet diet, this may limit the possible micronutrient malnutrition of poorly planned vegetarian and vegan diets (12). A single cohort study with a large proportion of vegetarian participants from the UK reported lower risk of type 2 diabetes with greater adherence to the EAT-Lancet diet (13), but further research in omnivorous dietary contexts is needed. Therefore, we investigated the association between adherence to the EAT-Lancet diet and incident type 2 diabetes in a Danish population of middle-aged adults.

## Research Design and Methods

### Study population and design

The Danish Diet, Cancer and Health cohort comprises 57,053 participants. To be eligible for participation in the cohort individuals had to be born in Denmark, live in Copenhagen or Aarhus County, be 50-64 years old, and have no previous diagnosis of cancer in the Danish Cancer Registry. Of the 160,725 eligible persons invited during the recruitment period in 1993-1997, 57,053 participated in the study (14). Participants completed questionnaires on diet and lifestyle and visited one of two study centres for anthropometric and other biological measurements. Questionnaires were optically scanned at the study centre to check for errors and missing information. Afterwards, a lab technician clarified all unclear information with participants (15, 16). Relevant ethics committees and the Danish Data Protection Agency approved the study. All participants gave written informed consent.

Participants with cancer or diabetes before baseline, missing or incomplete dietary data, or missing data on covariates were excluded (Supplemental Figure 1).

### Assessment of the EAT-Lancet diet score

Prior to the study centre visit, dietary data were collected using a validated 192-item self-administered food frequency questionnaire (FFQ). Participants reported their average intake of different food and beverage items over the past 12 months within 12 possible categories, ranging from “never” to “eight times or more per day”. After applying sex-specific portion sizes, intake was estimated using the Danish national food composition database and a specifically designed software program (17, 18). The FFQ was validated against two diet records of seven consecutive days and found to be a valid instrument for measuring dietary intake over the past year (19).

This study used the 14 dietary components of the EAT-Lancet diet score constructed previously (13, 20) (Table 1). Each component contributed 0 (non-adherence) or 1 (adherence) point, resulting in a total score ranging 0-14 points. Adherence was categorized as 0-7 points (reference group), 8 points, 9 points, 10 points, and 11-14 points to ensure adequate numbers of cases in each group.

**Table 1.** EAT-Lancet scoring criteria. EAT-Lancet diet scoring criteria based on components from the Danish Diet, Cancer, and Health cohort (DCH) food frequency questionnaire (FFQ).

| **Dietary component** | **Dietary components from the DCH FFQ** | **Criteria for scoring 1 point** | **% (n) scoring 1 point (n=54,232)** |
| --- | --- | --- | --- |
| 1. Grains | Whole grains, refined grains | $\leq 464$ g/day, whole grain fibre $> 5$ g | 99.73 (54,083) |
| 2. Tubers and starchy vegetables | Potatoes, fatty potatoes | $\leq 100$ g/day | 34.27 (18,584) |
| 3. Vegetables | Leafy, stalk, root, and fruiting vegetables, cabbages, mushrooms, onion/garlic | $\geq 200$ g/day | 32.81 (17,793) |
| 4. Fruits | Citrus fruits, other fruits | $\geq 100$ g/day | 65.30 (35,416) |
| 5. Dairy foods | Skimmed milk, semi-skimmed milk, whole milk, buttermilk, cheese, whole-fat fermented, low-fat fermented | $\leq 500$ g/day | 70.77 (38,582) |
| 6. Beef, lamb, pork | Red meat, processed meat | $\leq 28$ g/day | 1.98 (1075) |
| 7. Chicken, other poultry | Poultry | $\leq 58$ g/day | 94.95 (51,491) |
| 8. Eggs | Eggs | $\leq 25$ g/day | 57.25 (31,047) |
| 9. Fish | Fresh and processed lean, medium-fat and fatty fish | $\leq 100$ g/day | 96.59 (52,385) |
| 10. Dry beans, lentils, peas | Legumes | $\leq 100$ g/day* | 100 (54,232) |
| 11. Soy foods | Soy | $\leq 50$ g/day | 100 (54,232) |
| 12. Peanuts or tree nuts | Nuts | $\geq 25$ g/day | 0.39 (210) |
| 13. Added fats | Saturated fatty acids, monounsaturated fatty acids,<br>polyunsaturated fatty acids | Ratio of 0.8 for<br>unsaturated: saturated fat<br>intake | 99.37 (53,893) |
| 14. Added sugars | Added sugar | $\leq 31$ g/day | 51.61 (27,988) |

\* Dried, raw weight

### Outcomes

Incident type 2 diabetes cases were identified through register linkage to the Danish National Diabetes Registry using each participant’s unique national personal identification number. We defined incidence as the date of participants’ fulfilment of any of the following criteria: diagnosis in the Danish National Patient Registry; registration with chiropody (as a diabetic patient) in the National Health Service Registry; five blood-glucose measurements in a 1-y period or two blood-glucose measures per year in five consecutive years in the National Health Service Registry; purchased oral antidiabetic drugs in the Danish National Prescription Registry; or purchased prescribed insulin recorded in the National Health Service Registry (21).

The Danish National Diabetes Registry was validated against medical records with a positive predictive value of 80% (21, 22). The registry was incomplete until January 1^st^ 1995, but included prevalent cases from January 1^st^ 1990. Therefore, information on incident cases before January 1^st^ 1995 was incomplete and baseline was set to January 1^st^ 1995 for those recruited before that date (n=6) (21, 23). Diabetes cases in the Danish National Diabetes Registry were not differentiated between type 1 and type 2 diabetes (22), but as participants were middle-aged at baseline, all incident diabetes cases are assumed to be of type 2 diabetes.

Vital status, as well as emigration dates, were obtained through linkage using participants’ national personal identification numbers to The Danish Civil Registration System (24).

### Covariates

Data on lifestyle habits were collected through a self-administered lifestyle questionnaire and included questions on sex, smoking history, educational level, physical activity, and previous history of hypertension and hypercholesterolemia. Information on alcohol intake was assessed through the FFQ (15, 16).

Height and weight were measured by trained personnel to the closest 0.5 cm and 0.1 kg respectively at a study center. Body mass index was calculated as weight divided by height squared (kg/m^2^) (14). Waist circumference was measured by trained personnel to the closest 0.5 cm at the natural waist or midway between lowest rib and iliac crest with a non-stretchable measuring tape (14).

### Statistical analyses

Standard summary statistics were used to describe participant characteristics across categories of EAT-Lancet diet scores.

Cox proportional hazards models with age as underlying time scale were used to estimate the association between EAT-Lancet diet score and risk of type 2 diabetes. Participants were considered at risk from entry to the study to censoring, which was the date of type 2 diabetes diagnosis, death, emigration, or December 31^st^ 2011; whichever came first. The results are presented as hazard ratios (HR) with 95% confidence intervals (CI).

The analysis included four levels of adjustment. Covariates were selected *a priori* based on a review of the literature and directed acyclic graphs (Supplemental Figure 2). Model 1a adjusted for age. Model 1b further adjusted for physical activity (≥30 min/day, <30 min/day of moderate-to-vigorous physical activity), education (vocational; short 1-2 years; medium 3-4 years; high >4 years), smoking status (never; former; current <15 g tobacco/day; current 15–25 g tobacco/day; current >25 g tobacco/day), alcohol intake (g/day; continuous, as restricted cubic splines with 5 knots), and sex. Because adiposity, hypertension, and hypercholesterolemia may act as mediators rather than confounders, model 2 further adjusted for history of hypertension (yes; no; do not know), history of hypercholesterolemia (yes; no; do not know), waist circumference (cm; continuous), and body mass index (kg/m^2^; continuous, as restricted cubic splines with 4 knots). Because high EAT-Lancet scores are achievable with different energy intakes, we further adjusted model 2 for energy intake (kJ/day, continuous) (model 3) to account for variation in total energy intake.

In sensitivity analyses, we stratified the study population by median energy intake and sex to assess the association between EAT-Lancet diet score and risk of type 2 diabetes across the different strata. We also investigated different cut-points for the EAT-Lancet diet score to assess robustness. To estimate the minimum strength of association that an unmeasured confounder would need to have to explain away the observed associations, conditioned on measured covariates, E-values were calculated (25).

## Results

The median follow-up time was 15 years and 4 months, and 7130 cases of type 2 diabetes (3959 in men, and 3171 in women) were identified.

Participants scoring 11-14 points were more likely to be women, have a longer education, be non-smokers, and report a history of hypertension or hypercholesterolemia than those scoring 0-7 points (Table 2). After adjustment for potential confounders (Table 3, model 1b) a 22% (95% CI: 14%; 29%) lower risk of type 2 diabetes was observed among those scoring 11-14 points compared with those scoring 0-7 points. When further adjusting for potential mediators, results were similar in magnitude to model 1b. Adjusting for total energy intake, and thus comparing participants scoring 11-14 points compared with those scoring 0-7 points for a given energy intake, showed a 19% (95% CI: 27%;11%) lower risk of type 2 diabetes (model 3).

**Table 2.** Baseline characteristics of the Danish Diet, Cancer, and Health cohort participants eligible for analysis across EAT-Lancet diet scores

| EAT-Lancet diet score | Total population<br>n= 54,242 | 0-7 points<br>n= 6271 | 8 points<br>n= 12,074 | 9 points<br>n= 16,235 | 10 points<br>n= 12,404 | 11-14 points<br>n= 7248 |
| --- | --- | --- | --- | --- | --- | --- |
| Female (% , n) | 52.6, 28,505 | 30.9, 1939 | 41.2, 4968 | 51.2, 8318 | 63.3, 7856 | 74.8, 5424 |
| Age (median years, 5-95% P*) | 56, 50-64 | 56, 50-64 | 56, 50-64 | 56, 50-64 | 55, 50-64 | 55, 50-63 |
| Educational duration (% , n) |  |  |  |  |  |  |
| Elementary school | 14.7, 7959 | 18.5, 1161 | 16.7, 2020 | 14.9, 2418 | 12.9, 1605 | 10.4, 755 |
| Short | 23.0, 12,450 | 20.2, 1266 | 21.4, 2581 | 22.9, 3721 | 24.0, 2979 | 26.3, 1903 |
| Medium | 40.2, 21,776 | 40.8, 2560 | 40.3, 4860 | 39.8, 6458 | 40.4, 5011 | 39.8, 2887 |
| Long | 22.2, 12,047 | 20.5, 1284 | 21.6, 2613 | 22.4, 3638 | 22.7, 2809 | 23.5, 1703 |
| Smokers (% , n) | 35.9, 19,493 | 46.6, 2923 | 40.9, 4935 | 36.6, 5935 | 31.2, 3870 | 25.3, 1830 |
| BMI (mean kg/m<sup>2</sup> ± SD) | 26.0 ± 4.0 | 26.4 ± 4.0 | 26.1 ± 4.0 | 26.0 ± 4.0 | 25.8 ± 4.0 | 25.5 ± 4.0 |
| Waist circumference (mean cm ± SD) |  |  |  |  |  |  |
| Females | 81.9 ± 11.1 | 83.07±12.00 | 82.34±11.24 | 82.22±11.00 | 81.67±10.97 | 80.92±10.72 |
| Males | 95.9 ± 9.8 | 96.38±10.13 | 96.01±9.90 | 95.90±9.85 | 95.54±9.55 | 94.51±9.42 |
| Physical activity ≥ 30 min/day (% , n) | 39.6, 21,495 | 37.2, 2334 | 37.4, 4511 | 38.4, 6237 | 41.2, 5110 | 45.6, 3303 |
| Alcohol intake (median g/day, 5%-95% P*) | 13.0, 0.7-64.4 | 14.5, 0.5-71.1 | 13.7, 0.7-68.4 | 13.4, 0.8-65.5 | 12.6, 0.7-60.3 | 11.6, 0.6-52.8 |
| Energy intake (median MJ/day, 5-95% P*) | 8.9, 5.5-13.9 | 10.8, 7.3-16.6 | 9.7, 6.4-14.6 | 8.9, 5.6-13.4 | 8.2, 5.1-12.3 | 7.6, 4.9-11.0 |
| History of hypertension (% , n) | 15.7, 8491 | 13.5, 847 | 15.3, 1852 | 16.3, 2640 | 15.8, 1963 | 16.4, 1189 |
| History of hypercholesterolemia (% , n) | 7.2, 3911 | 5.9, 367 | 6.8, 821 | 7.5, 1214 | 7.9, 982 | 7.3, 527 |

\*Percentiles

**Table 3.** Age- and multivariable adjusted hazard ratios (HR) and 95% confidence intervals (CI) for incident type 2 diabetes based on EAT-Lancet diet score in the Danish Diet, Cancer and Health cohort.

| EAT-Lancet diet score | Events, *n* | IR* per 1000 person-years [95% CI] | Model 1a†<br>HR [95% CI] | Model 1b‡<br>HR [95% CI] | Model 2§<br>HR [95% CI] | Model 3\|\|<br>HR [95% CI] |
| --- | --- | --- | --- | --- | --- | --- |
| 0-7 | 1025 | 12.00 [11.29; 12.76] | Reference | Reference | Reference | Reference |
| 8 | 1735 | 10.34 [9.87; 10.84] | 0.86 [0.79; 0.93] | 0.91 [0.84; 0.99] | 0.92 [0.85; 0.99] | 0.91 [0.84; 0.98] |
| 9 | 2173 | 9.52 [9.13; 9.93] | 0.80 [0.74; 0.86] | 0.90 [0.84; 0.98] | 0.91 [0.85; 0.98] | 0.89 [0.83; 0.97] |
| 10 | 1463 | 8.21 [7.84; 8.64] | 0.69 [0.64; 0.75] | 0.84 [0.77; 0.92] | 0.85 [0.79; 0.93] | 0.83 [0.76; 0.90] |
| 11-14 | 734 | 6.94 [6.46; 7.46] | 0.59 [0.54; 0.65] | 0.78 [0.71; 0.86] | 0.83 [0.76; 0.92] | 0.81 [0.73; 0.89] |

\* Incidence rate
† Adjusted for age
‡ Further adjusted for physical activity ( $\geq 30$ min/day, $< 30$ min/day of moderate-to-vigorous physical activity), education (elementary school; short 1-2 years; medium 3-4 years; high $> 4$ years), smoking status (never; former; current $< 15$ g tobacco/day; current 15–25 g tobacco/day; current $> 25$ g tobacco/day), alcohol intake (g/day; restricted cubic splines with 5 knots), and sex.
§ Further adjusted for history of hypertension (yes, no, do not know), history of hypercholesterolemia (yes, no, do not know), waist circumference (cm; continuous), and body mass index ( $\text{kg}/\text{m}^2$ ; continuous, as restricted cubic splines with 5 knots).
|| Further adjusted for energy intake (MJ/day; continuous).

### Sensitivity analyses

When stratifying the population by median energy intake we found in model 1b, for both strata, similar associations between EAT-Lancet diet and risk of type 2 diabetes as in the main analysis (Table 4). Stratification by sex (Table 4) showed lower relative risk of developing type 2 diabetes with higher EAT-Lancet adherence for men than for women (Men HR: 0.68, 95% CI: 0.58; 0.79; Women HR: 0.83, 95% CI: 0.71; 0.96). When altering the cut-points for the EAT-Lancet diet score to 0-7; 8; 9; 10-14, or to cut-points used in a previous study (13), results showed similar patterns of lower risk of type 2 diabetes with greater adherence (Supplemental Table 1 and 2). The E-value calculations (Supplemental Table 3) indicated that to explain away the association between EAT-Lancet diet scores and type 2 diabetes (model 1b, comparing the highest to the lowest level of adherence), the minimum strength of association between an unmeasured confounder and EAT-Lancet score and type 2 diabetes had to be a risk ratio of 1.66. For model 2, the association had to be 1.53, and for model 3, 1.58.

**Table 4.** Association between EAT-Lancet diet score and risk of type 2 diabetes across stratifications

| EAT-Lancet score | 0-7 | 8 | 9 | 10 | 11-14 |
| --- | --- | --- | --- | --- | --- |
| Sex<sup>*</sup> |  |  |  |  |  |
| Male | Reference | 0.88 [0.80; 0.96] | 0.86 [0.78; 0.94] | 0.78 [0.71; 0.87] | 0.68 [0.58; 0.79] |
| Female | Reference | 0.98 [0.84; 1.13] | 0.96 [0.83; 1.10] | 0.88 [0.77; 1.02] | 0.83 [0.71; 0.96] |
| Energy intake<sup>†</sup> |  |  |  |  |  |
| <8896 kJ/day | Reference | 0.88 [0.75; 1.03] | 0.90 [0.78; 1.04] | 0.83 [0.71; 0.96] | 0.72 [0.61; 0.85] |
| ≥8896 kJ/day | Reference | 0.90 [0.82; 0.98] | 0.83 [0.76; 0.92] | 0.74 [0.66; 0.83] | 0.72 [0.61; 0.84] |

\* Adjusted for age, physical activity ( $\geq 30$ min/day, <30 min/day of moderate-to-vigorous physical activity), education (elementary school; short 1-2 years; medium 3-4 years; high >4 years), smoking status (never; former; current <15 g tobacco/day; current 15–25 g tobacco/day; current >25 g tobacco/day), alcohol intake (g/day; restricted cubic splines with 5 knots).
† Adjusted for age, physical activity ( $\geq 30$ min/day, <30 min/day of moderate-to-vigorous physical activity), education (elementary school; short 1-2 years; medium 3-4 years; high >4 years), smoking status (never; former; current <15 g tobacco/day; current 15–25 g tobacco/day; current >25 g tobacco/day), alcohol intake (g/day; restricted cubic splines with 5 knots), and sex.

## Conclusions

In this cohort of Danish middle-aged adults, greater adherence to the EAT-Lancet diet was associated with a lower risk of type 2 diabetes. These findings suggest a public health benefit concerning type 2 diabetes prevention when following this dietary pattern.

This study has several strengths, such as a large and representative sample size with a long follow-up period, minimal loss to follow-up (0.7%) and outcome data obtained from a nation-wide registry. The negligible loss to follow-up ensures very low risk of selection bias of our results. A limitation is potential measurement errors in diet assessment using a FFQ that could lead to bias towards the null due to non-differential misclassification of the exposure. A single measure of diet is unable to capture changes in diet over time, which could also lead to misclassification of the exposure during follow-up. The EAT-Lancet diet score only included two levels of adherence for each component: adherence or non-adherence. The crude scoring will categorize participants together who are almost adherent with those who are far from adhering. For instance, 90% adherence in each component would nevertheless score 0 on total EAT-Lancet adherence, which a more graduated score could have accounted for. This might underestimate the association between EAT-Lancet diet adherence and type 2 diabetes, and pose a particular problem in the middle adherence categories, where scores are undifferentiated. However, this score has been used previously (13) and the categorizations used in our study led to results that were robust in sensitivity analyses with alternative groupings. The simple scoring system may have benefits in communicating our results to the public, as individuals can easily assess their own adherence to each component. Adjustment for energy intake allowed comparison of EAT-Lancet scores for a given total energy intake, which, as the diet can in principle be achieved at very low total energy intakes, ensured that variation in total energy intake did not explain our results. The stratified analysis showed similar associations between EAT-Lancet diet and type 2 diabetes in the two strata of energy intake. Stratification on sex showed a somewhat more beneficial association between EAT-Lancet score and risk of type 2 diabetes for men compared to women. Women are more likely to misreport dietary habits in questionnaires than men (26-28), which can cause greater misclassification of the exposure among women compared to men. This misclassification would most likely be non-differential and bias the estimated association toward the null. Thus, the true association between EAT-Lancet score and risk of type 2 diabetes might therefore be greater for women than what we observed in our study (26-28).

### Potential confounding

Information on smoking, alcohol consumption and physical activity are possibly error prone, for example due to social desirability bias. We adjusted for several variables that markedly changed the estimated association, and considerable confounding would have to remain to explain our results, although residual confounding cannot be completely excluded. Data collection provided no information on sleep or stress, both known to affect dietary patterns and risk of type 2 diabetes (29-33). However, E-values for our results indicate that the relative risk association between unmeasured confounders, such as sleep and stress, and diet or incident type 2 diabetes would each have to be 1.66 to explain away our results. A systematic review and meta-analysis has indicated that short sleep duration compared to 7-8 hours of sleep was associated with a higher risk of type 2 diabetes (RR: 1.28, 95% CI: 1.03-1.60). During a 9-year follow-up period high cortisol levels were also related to higher odds of type 2 diabetes onset compared to normal cortisol levels (OR: 1.18, 95% CI: 1.01-1.37). Our results may thus be residually confounded by stress and sleep, but these factors most likely do not account for the total estimated association.

### Other research and potential mechanisms

Other cohort studies have suggested that meat-restricted diets are associated with lower risk of type 2 diabetes, which is consistent with the results from this study (4, 13). Greater adherence to the EAT-Lancet diet has been hypothesized to have similar co-benefits regarding incident type 2 diabetes and environmental impact as vegan or vegetarian diets (12). As our study included individuals with an omnivorous dietary pattern, the results indicate that complete exclusion of meats or animal products is not necessary to achieve beneficial effects in relation to prevention of type 2 diabetes. The EAT-Lancet diet may be easier to adopt for most meat-preferring populations, such as in many Western countries, since the dietary pattern, despite a reduction in animal products, is somewhat similar to the general omnivorous dietary pattern. Recently, a modelling study concluded that adoption of the EAT-Lancet diet would be associated with a 13 % reduction in greenhouse gas emissions compared to adherence to national dietary guidelines, on average (34). Shifting diets towards the EAT-Lancet diet may therefore promote population health and lower greenhouse gas emissions simultaneously.

Plausible mechanisms underlying our results may be the high intake of dietary fibres resulting in a feeling of satiety and a stabilizing effect on blood glucose (15). The lower risk of type 2 diabetes associated with consumption of less red meat daily might be due to the concomitant higher consumption of plant-based foods, a replacement effect previously linked to lower risk of type 2 diabetes (4). In our study, participants with a high EAT-Lancet score were more likely to be women (Table 2). This may present challenges for accept of and adherence to dietary guidelines based on the EAT-Lancet diet for men particularly. Men are further from adherence to the EAT-Lancet diet due to e.g. a high consumption of meats compared to women in the general society (35). Thus, promotion of the EAT-Lancet diet and dietary recommendations based on this diet could be targeted men specifically to increase compliance.

### Conclusion and implications

This cohort study found that adherence to the EAT-Lancet diet was associated with a lower risk of incident type 2 diabetes in a middle-aged Danish population. Future cohort studies would benefit from focusing on refining the EAT-Lancet diet score and evaluating long-term consequences in other dietary contexts (e.g. low- and middle-income countries) whereas trials could focus on adaptation to the diet and on shorter-term physiological implications of adherence.

## Data Availability

This study was based on data from the Diet, Cancer and Health Cohort and are not publicly available due to personal information content, but can be acquired upon reasonable request from the Danish Cancer Society.

## Author contributions

The Danish Diet, Cancer and Health cohort was initiated by KO and AT as principal investigators. CCD, DBI, FL and AO conceived the research question. CCD, FL and DBI designed the analysis plan. FL and DBI did the data analysis. FL, DBI and CCD drafted the manuscript. All authors interpreted the results and critically revised the article for important intellectual content and gave final approval of the version to publish. CCD takes responsibility for the contents of this article.

The authors acknowledge the participants that provided data and the members of the study team that collected the data. The authors thank Lone Fredslund for assistance on data management.

## References

1. Saeedi P, Petersohn I, Salpea P, Malanda B, Karuranga S, Unwin N, Colagiuru S, Guariguata L, Motala AA, Ogurtsova K, Shaw JE, Bright D, Williams R. Global and regional diabetes prevalence estimates for 2019 and projections for 2030 and 2045: Results from the International Diabetes Federation Diabetes Atlas, 9th edition. Diabetes Research and Clinical Practice. 2019;157. doi: 10.1016/j.diabres.2019.107843. PubMed PMID: 107843.

2. Khan MAB, Hashim MJ, King JK, Govender RD, Mustafa H, Al Kaabi J. Epidemiology of Type 2 Diabetes - Global Burden of Disease and Forecasted Trends. J Epidemiol Glob Health. 2020;10(1):107–11. Epub 2020/03/17. doi: 10.2991/jegh.k.191028.001. PubMed PMID: 32175717; PubMed Central PMCID: PMC7310804.

3. Lin X, Xu Y, Pan X, Xu J, Ding Y, Sun X, Song X, Ren Y, Shan P. Global, regional, and national burden and trend of diabetes in 195 countries and territories: an analysis from 1990 to 2025. Scientific Reports. 2020;10(1):14790. doi: 10.1038/s41598-020-71908-9.

4. Qian F, Liu G, Hu FB, Bhupathiraju SN, Sun Q. Association Between Plant-Based Dietary Patterns and Risk of Type 2 Diabetes: A Systematic Review and Meta-analysis. JAMA Intern Med. 2019;179(10):1335–44. Epub 2019/07/23. doi: 10.1001/jamainternmed.2019.2195. PubMed PMID: 31329220; PubMed Central PMCID: PMC6646993

5. Hu FB. Plant-based foods and prevention of cardiovascular disease: an overview. The American Journal of Clinical Nutrition. 2003;78(3):544S–51S. doi: 10.1093/ajcn/78.3.544S.

6. Kahleova H, Levin S, Barnard N. Cardio-Metabolic Benefits of Plant-Based Diets. Nutrients. 2017;9(8). Epub 2017/08/10. doi: 10.3390/nu9080848. PubMed PMID: 28792455; PubMed Central PMCID: PMC5579641.

7. Kahleova H, Pelikanova T. Vegetarian Diets in the Prevention and Treatment of Type 2 Diabetes. J Am Coll Nutr. 2015;34(5):448–58. Epub 2015/04/29. doi: 10.1080/07315724.2014.976890. PubMed PMID: 25915002.

8. Gijsbers L, Ding EL, Malik VS, de Goede J, Geleijnse JM, Soedamah-Muthu SS. Consumption of dairy foods and diabetes incidence: a dose-response meta-analysis of observational studies. The American Journal of Clinical Nutrition. 2016;103(4):1111–24. doi: 10.3945/ajcn.115.123216.

9. Zhang M, Picard-Deland E, Marette A. Fish and Marine Omega-3 Polyunsatured Fatty Acid Consumption and Incidence of Type 2 Diabetes: A Systematic Review and Meta-Analysis. International Journal of Endocrinology. 2013;2013:501015. doi: 10.1155/2013/501015.

10. Mozaffarian D. Dietary and Policy Priorities for Cardiovascular Disease, Diabetes, and Obesity. Circulation. 2016;133(2):187–225. doi: 10.1161/CIRCULATIONAHA.115.018585.

11. Hemler EC, Hu FB. Plant-Based Diets for Cardiovascular Disease Prevention: All Plant Foods Are Not Created Equal. Current Atherosclerosis Reports. 2019;21(5):18. doi: 10.1007/s11883-019-0779-5.

12. Willett W, Rockström J, Loken B, Springmann M, Lang T, Vermeulen S, Garnett T, Tilman D, DeClerck F, Wood A, Jonell M, Clark M, Gordon LJ, Fanzo J, Hawkes C, Zurayk R, Rivera JA, De Vries W, Sibanda ML, Afshin A, Chaudhary A, Herrero M, Agustina R, Branca F, Lartey A, Fan S, Crona B, Fox E, Bignet V, Troell M, Lindahl T, Singh S, Cornell SE, Reddy KS, Narain S, Nishtar S, Murray CJL. Food in the Anthropocene: the EAT-Lancet Commission on healthy diets from sustainable food systems. Lancet. 2019;393(10170):447–92. Epub 2019/01/21. doi: 10.1016/s0140-6736(18)31788-4. PubMed PMID: 30660336.

13. Knuppel A, Papier K, Key TJ, Travis RC. EAT-Lancet score and major health outcomes: the EPIC-Oxford study. Lancet. 2019;394(10194):213–4. Epub 2019/06/27. doi: 10.1016/s0140-6736(19)31236-x. PubMed PMID: 31235280.

14. Tjønneland A, Olsen A, Boll K, Stripp C, Christensen J, Engholm G, Overvad K. Study design, exposure variables, and socioeconomic determinants of participation in Diet, Cancer and Health: a population-based prospective cohort study of 57,053 men and women in Denmark. Scand J Public Health. 2007;35(4):432–41. Epub 2007/09/06. doi: 10.1080/14034940601047986. PubMed PMID: 17786808.

15. Kyrø C, Tjønneland A, Overvad K, Olsen A, Landberg R. Higher Whole-Grain Intake Is Associated with Lower Risk of Type 2 Diabetes among Middle-Aged Men and Women: The Danish Diet, Cancer, and Health Cohort. The Journal of Nutrition. 2018;148(9):1434–44. doi: https://doi.org/10.1093/jn/nxy112.

16. Andersen JLM, Halkjær J, Rostgaard-Hansen AL, Martinussen N, Lund AQ, Kyrø C, et al. Intake of whole grain and associations with lifestyle and demographics: a cross-sectional study based on the Danish Diet, Cancer and Health-Next Generations cohort. Eur J Nutr. 2021;60(2):883–95. Epub 2020/06/06. doi: 10.1007/s00394-020-02289-y. PubMed PMID: 32500314.

17. DTU Food. Danish National Food Tables 2019 [cited 2021 October 7]. Available from: https://frida.fooddata.dk/?lang=en.

18. Lauritsen A. FoodCalc 2019 [cited 2021 October 7]. Available from: https://www.cancer.dk/dchdata/access-to-data-and-biobank/foodcalc/.

19. Tjønneland A, Overvad K, Haraldsdóttir J, Bang S, Ewertz M, Jensen OM. Validation of a semiquantitative food frequency questionnaire developed in Denmark. Int J Epidemiol. 1991;20(4):906–12. Epub 1991/12/01. doi: 10.1093/ije/20.4.906. PubMed PMID: 1800429.

20. Knuppel A, Papier K, Key TJ, Travis RC. Supplement to: EAT-Lancet score and major health outcomes: the EPIC-Oxford study. Lancet. 2019;394(10194):213–4. Epub 2019/06/27. doi: 10.1016/s0140-6736(19)31236-x. PubMed PMID: 31235280.

21. Carstensen B, Kristensen JK, Marcussen MM, Borch-Johnsen K. The National Diabetes Register. Scand J Public Health. 2011;39(7 Suppl):58–61. Epub 2011/08/04. doi: 10.1177/1403494811404278. PubMed PMID: 21775353.

22. Green A, Sortsø C, Jensen PB, Emneus M. Validation of the danish national diabetes register. Clin Epidemiol. 2014;7:5–15. doi: 10.2147/CLEP.S72768. PubMed PMID: 25565889.

23. Sørensen M, Andersen ZJ, Nordsborg RB, Becker T, Tjønneland A, Overvad K, Raaschou-Nielsen O. Long-term exposure to road traffic noise and incident diabetes: a cohort study. Environ Health Perspect. 2013;121(2):217–22. Epub 2012/12/12. doi: 10.1289/ehp.1205503. PubMed PMID: 23229017; PubMed Central PMCID: PMC3569689.

24. Pedersen CB. The Danish Civil Registration System. Scand J Public Health. 2011;39(7 Suppl):22–5. doi: 10.1177/1403494810387965. PubMed PMID: 21775345.

25. VanderWeele TJ, Ding, P. Sensitivity Analysis in Observational Research: Introducing the E-Value. Annals of Internal Medicine. 2017;167(4):268–74. doi: 10.7326/m16-2607%m28693043.

26. Schatzkin A, Kipnis V, Carroll RJ, Midthune D, Subar AF, Bingham S, Schoeller DA, Troiano RP, Freedman LS. A comparison of a food frequency questionnaire with a 24-hour recall for use in an epidemiological cohort study: results from the biomarker-based Observing Protein and Energy Nutrition (OPEN) study. Int J Epidemiol. 2003;32(6):1054–62. Epub 2003/12/19. doi: 10.1093/ije/dyg264. PubMed PMID: 14681273.

27. Subar AF, Kipnis V, Troiano RP, Midthune D, Schoeller DA, Bingham S, Sharbaugh CO, Trabulsi J, Runswick S, Ballard-Barbash R, Sunshine J, Schatzkin A. Using intake biomarkers to evaluate the extent of dietary misreporting in a large sample of adults: the OPEN study. Am J Epidemiol. 2003;158(1):1–13. Epub 2003/07/02. doi: 10.1093/aje/kwg092. PubMed PMID: 12835280.

28. Kipnis V, Subar AF, Midthune D, Freedman LS, Ballard-Barbash R, Troiano RP, Bingham S, Schoeller DA, Schatzkin A, Carroll RJ. Structure of Dietary Measurement Error: Results of the OPEN Biomarker Study. American Journal of Epidemiology. 2003;158(1):14–21. doi: 10.1093/aje/kwg091.

29. Briançon-Marjollet A, Weiszenstein M, Henri M, Thomas A, Godin-Ribuot D, Polak J. The impact of sleep disorders on glucose metabolism: endocrine and molecular mechanisms. Diabetology & Metabolic Syndrome. 2015;7(1):25. doi: 10.1186/s13098-015-0018-3.

30. Chaput JP, Després JP, Bouchard C, Tremblay A. The association between short sleep duration and weight gain is dependent on disinhibited eating behavior in adults. Sleep. 2011;34(10):1291–7. Epub 2011/10/04. doi: 10.5665/sleep.1264. PubMed PMID: 21966060; PubMed Central PMCID: PMC3174831.

31. McNeil J, Doucet É, Chaput JP. Inadequate sleep as a contributor to obesity and type 2 diabetes. Can J Diabetes. 2013;37(2):103–8. Epub 2013/09/28. doi: 10.1016/j.jcjd.2013.02.060. PubMed PMID: 24070800.

32. Cappuccio FP, D’Elia L, Strazzullo P, Miller MA. Quantity and quality of sleep and incidence of type 2 diabetes: a systematic review and meta-analysis. Diabetes Care. 2010;33(2):414–20. Epub 2009/11/17. doi: 10.2337/dc09-1124. PubMed PMID: 19910503; PubMed Central PMCID: PMC2809295.

33. Joseph JJ, Golden SH. Cortisol dysregulation: the bidirectional link between stress, depression, and type 2 diabetes mellitus. Ann N Y Acad Sci. 2017;1391(1):20–34. Epub 2016/10/18. doi: 10.1111/nyas.13217. PubMed PMID: 27750377; PubMed Central PMCID: PMC5334212.

34. Springmann M, Spajic L, Clark MA, Poore J, Herforth A, Webb P, Rayner M, Scarborough P. The healthiness and sustainability of national and global food based dietary guidelines: modelling study. BMJ. 2020;370:m2322. doi: 10.1136/bmj.m2322.

35. Pedersen AN, Christensen T, Matthiessen J, Knudsen VK, Sørensen MR, Biltoft-Jensen AP, Hinsch H-J, Ygil KH, Kørup K, Saxholt E, Trolle E, Søndergaard AB, Fagt S. Dietary habits in Denmark 2011-2013. Main results. Copenhagen, Denmark: DTU Food, 2015.

