## Supplemental material for "Adherence to the EAT-Lancet diet and risk of incident type 2 diabetes: the Danish Diet, Cancer and Health cohort"

### Supplemental Table 1 – Sensitivity Analysis

*Supplemental Table 1. Risk of type 2 diabetes in the Danish Diet, Cancer and Health cohort (n=54,232) estimated on four cut-points of the EAT-Lancet diet score.*

| | Statistical model | Statistical model | Statistical model | Statistical model |
|---|---|---|---|---|
| EAT-Lancet adherence score | Model 1a* HR [95% CI] | Model 1b† HR [95% CI] | Model 2‡ HR [95% CI] | Model 3§ HR [95% CI] |
| 0-7 | Reference group | Reference group | Reference group | Reference group |
| 8 | 0.86 [0.79; 0.93] | 0.91 [0.85; 0.99] | 0.92 [0.85; 1.00] | 0.91 [0.84; 0.98] |
| 9 | 0.80 [0.74; 0.86] | 0.91 [0.84; 0.98] | 0.91 [0.85; 0.98] | 0.89 [0.83; 0.97] |
| 10-14 | 0.66 [0.61; 0.71] | 0.82 [0.76; 0.89] | 0.85 [0.79; 0.92] | 0.82 [0.76; 0.89] |

* Adjusted for age

†Further adjusted for physical activity (≥30 min/day, <30 min/day of moderate-to-vigorous physical activity), education (elementary school; short 1-2 years; medium 3-4 years; high >4 years), smoking status (never; former; current <15 g tobacco/day; current 15–25 g tobacco/day; current >25 g tobacco/day), alcohol intake (g/day; restricted cubic splines with 5 knots), and sex.

‡ Further adjusted for history of hypertension (yes, no, do not know), history of hypercholesterolemia (yes, no, do not know), waist circumference (cm; continuous), and body mass index (kg/m$^2$; continuous, as restricted cubic splines with 5 knots).

§ Further adjusted for energy intake (MJ/day; continuous).

### Supplemental Table 2 – Sensitivity Analysis

*Supplemental Table 2. Risk of type 2 diabetes in the Danish Diet, Cancer and Health cohort (n=54,232) estimated on cut-points of the EAT-Lancet dietary score from EPIC-Oxford (1).*

| | Statistical model | Statistical model | Statistical model | Statistical model |
|---|---|---|---|---|
| EAT-Lancet adherence score | Model 1a* HR [95% CI] | Model 1b† HR [95% CI] | Model 2‡ HR [95% CI] | Model 3§ HR [95% CI] |
| 0-9 | Reference group | Reference group | Reference group | Reference group |
| 10 | 0.81 [0.76; 0.86] | 0.91 [0.86; 0.97] | 0.92 [0.86; 0.97] | 0.91 [0.86; 0.97] |
| 11 | 0.72 [0.66; 0.79] | 0.87 [0.80; 0.95] | 0.92 [0.84; 1.00] | 0.90 [0.83; 0.99] |
| 12-14 | 0.58 [0.48; 0.69] | 0.73 [0.61; 0.88] | 0.82 [0.68; 0.98] | 0.81 [0.67; 0.97] |

* Adjusted for age

†Further adjusted for physical activity (≥30 min/day, <30 min/day of moderate-to-vigorous physical activity), education (elementary school; short 1-2 years; medium 3-4 years; high >4 years), smoking status (never; former; current <15 g tobacco/day; current 15–25 g tobacco/day; current >25 g tobacco/day), alcohol intake (g/day; restricted cubic splines with 5 knots), and sex.

‡ Further adjusted for history of hypertension (yes, no, do not know), history of hypercholesterolemia (yes, no, do not know), waist circumference (cm; continuous), and body mass index (kg/m$^2$; continuous, as restricted cubic splines with 5 knots).

§ Further adjusted for energy intake (MJ/day; continuous)

### Supplemental Table 3 – E-values

*Supplemental Table 3. E-values and 95% CI based on HRs from main analysis*

| | E-value [95% CI] | | |
|---|---|---|---|
| EAT-Lancet score | Model 1b* HR [95%CI] | Model 2† HR [95%CI] | Model 3‡ HR [95%CI] |
| 0-7 | Reference | Reference | Reference |
| 8 | 1.34 [1.09] | 1.31 [1.13] | 1.34 [1.13] |
| 9 | 1.36 [1.13] | 1.34 [1.13] | 1.39 [1.17] |
| 10 | 1.51 [1.31] | 1.48 [1.28] | 1.53 [1.36] |
| 11-14 | 1.66 [1.46 ] | 1.53 [1.31] | 1.58 [1.39] |

*Adjusted for age (years) physical activity (≥30 min/day, <30 min/day of moderate-to-vigorous physical activity), education (elementary school; short 1-2 years; medium 3-4 years; high >4 years), smoking status (never; former; current <15 g tobacco/day; current 15–25 g tobacco/day; current >25 g tobacco/day), alcohol intake (g/day; restricted cubic splines with 5 knots), and sex.

† Further adjusted for history of hypertension (yes, no, do not know), history of hypercholesterolemia (yes, no, do not know), waist circumference (cm; continuous), and body mass index (kg/$m^2$; continuous, as restricted cubic splines with 5 knots).

‡ Further adjusted for energy intake (MJ/day; continuous)

**Supplemental Figure 1 – Flow diagram of study population**

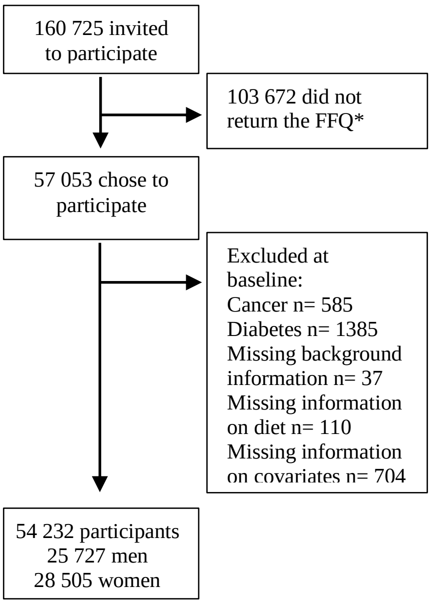

*Supplemental Figure 1. Flow diagram of the study population from the Danish Diet, Cancer and Health cohort who were eligible for statistical analysis. *FFQ, Food frequency questionnaire*

### Supplemental Figure 2 – Directed Acyclic Graphs

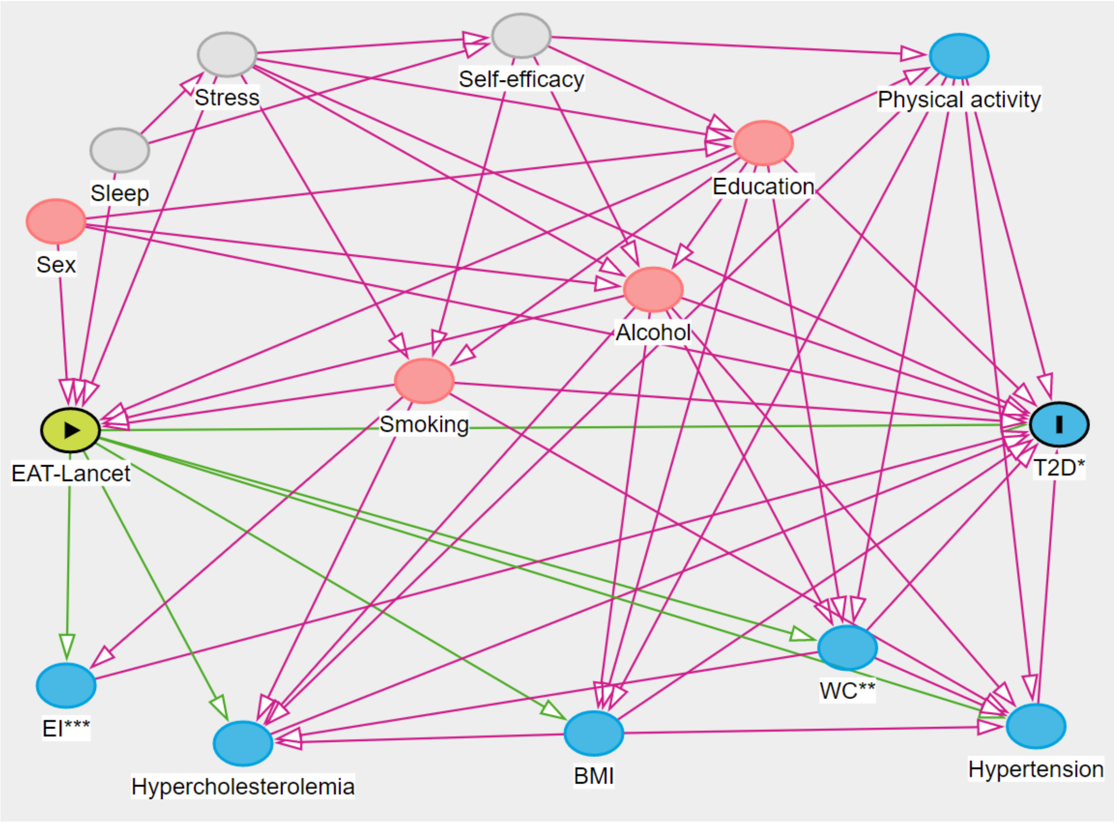

*Supplemental Figure 2. Directed Acyclic Graph of a priori presumed confounders before adjustment. This DAG was constructed in DAGitty (2). *T2D, type 2 diabetes; **WC, waist circumference; ***EI, energy intake; exposure; outcome; ancestor of outcome; ancestor of exposure and outcome; unobserved (latent); causal path; biasing path.*

### References

1. Knuppel A, Papier K, Key TJ, Travis RC. EAT-Lancet score and major health outcomes: the EPIC-Oxford study. Lancet. 2019;394(10194):213-4.

2. Textor J, van der Zander, B., Gilthorpe, M. K., Liskiewicz, M., Ellison, G. T. H. Robust causal inference using directed acyclic graphs: the R package 'dagitty'. International Journal of Epidemiology. 2016;45(6):1887-94.
